# Dynamics of antibody responses, cellular immunity, and breakthrough infections among Japanese healthcare workers during the 6 months after receiving two doses of BNT162b2 mRNA vaccine

**DOI:** 10.1101/2022.01.29.22270052

**Authors:** Yoshifumi Uwamino, Toshinobu Kurafuji, Kumiko Takato, Akiko Sakai, Akiko Tanabe, Masayo Noguchi, Yoko Yatabe, Tomoko Arai, Akemi Ohno, Yukari Tomita, Ayako Shibata, Hiromitsu Yokota, Wakako Yamasawa, Ho Namkoong, Yasunori Sato, Naoki Hasegawa, Masatoshi Wakui, Mitsuru Murata, the Keio Donner Project Team

**Author notes:** Corresponding author: Yoshifumi Uwamino, MD, PhD, Department of Laboratory Medicine, Keio University School of Medicine, 35 Shinanomachi, Shinjuku-ku, Tokyo 160-8582, Japan.

## Abstract

**Introduction:** The waning of the antibody titre after the first two doses of the Pfizer-BioNTech BNT162b2 mRNA SARS-CoV-2 vaccine was reported. However, knowledge of the dynamics of cellular immunity is scarce. Here, we performed a prospective cohort study to disclose antibody and cellular immunity dynamics and discuss the relationship between immunity and breakthrough infection.

**Methods:** The study had a prospective cohort design. Antibody titres against SARS-CoV-2 in serially collected serum samples of 608 Japanese vaccinees after 6 months of vaccination were measured. Simultaneously, T-cell immunity dynamics were assessed using the QuantiFERON SARS-CoV-2 assay. Additionally, participants with suspected breakthrough infection were detected according to the positive conversion of the IgG assay for nucleocapsid proteins of SARS-CoV-2.

**Results:** Antibody titres were elevated 3 weeks after vaccination and waned over the remainder of the study period. The QuantiFERON SARS-CoV-2 assay performed on 536 participants demonstrated the similar dynamics. Six participants without predisposing medical conditions demonstrated positive conversion of the IgG assay for nucleocapsid proteins, while five were asymptomatic.

**Conclusion:** Waning of humoral and cellular immunity within 6 months of administration of two doses of BNT162b2 vaccine among Japanese healthcare professionals and the occurrence of asymptomatic breakthrough infection was suspected in approximately 1 of 100 vaccinees.

(UMIN000043340)

## Introduction

Although 2 years have passed since the first cases of severe acute respiratory syndrome coronavirus 2 (SARS-CoV-2) infection were reported, the pandemic is ongoing. It was hoped that vaccination would end the pandemic; however, waning of the antibody titre after the first two doses of vaccination was reported [1] and cases of breakthrough infection have occurred [2]. The Pfizer-BioNTech BNT162b2 mRNA SARS-CoV-2 vaccine is a messenger RNA vaccine that is widely used in Japan and worldwide. Most large cohort studies describing antibody dynamics after two doses of BNT162b2 vaccine were conducted in Western countries. However, the immune response may be affected by racial and demographic factors. Although reports about declining immune response or cellular immunity during the time period among Japanese vaccinees are available, the sample size was limited [3–5]. We launched a prospective cohort study of more than 600 Japanese healthcare workers before the administration of the first dose of the BNT162b2 vaccine to observe humoral and cellular immune responses to the vaccine among Japanese people. We previously reported humoral and cellular responses immediately after two doses of BNT162b2 vaccine and its relationship with adverse reactions [6,7]. Here, we present the dynamics of antibody titre and cellular immunity during the 6 months after the initial two doses of BNT162b2 vaccine. Additionally, we describe cases in which breakthrough infections were suspected based on the positive seroconversion for SARS-CoV-2 nucleocapsid proteins.

## Participants and Methods

### Participants

Healthcare workers working on the Keio University Shinanomachi Campus (Tokyo, Japan) who were vaccinated against SARS-CoV-2 between 16 February and 9 March, 2021 were recruited for the study. The campus has a university hospital with 960 beds and a medical school. Written informed consent was obtained from all participants before mass vaccination. The study was approved by the ethics committee of the Keio University School of Medicine (approval no. 20200330). Mass vaccination was carried out using the BNT162b2 vaccine (Comirnaty intramuscular injection, Pfizer, New York, USA), which were stored and prepared according to the instructions given in the package insert. Each participant received two doses of vaccine, administered 3 weeks apart.

### Sample collection

Serial serum samples were collected from each participant at five time points. The first sampling point was before or on the same day as the first vaccination. The second sampling point was between 15 and 28 April, 2021, approximately 3 weeks after the second dose. The third sampling point was between 20 May and 2 June, 2021, approximately 8 weeks after the second dose. The fourth sampling point was between 28 June and 9 July, 2021, approximately 3 months after the second dose. The final sampling point was between 28 September and 8 October, 2021, approximately 6 months after the second dose. Answers to the questionnaires were obtained from each participant at the time of sampling. At the first sampling point, an inquiry into age, sex, height, body weight, use of systemic steroids or other immunosuppressants, ongoing cancer chemotherapy, and history of immunodeficiency, cancers, autoimmune diseases, diabetes, and COVID-19 was made. At the second to final sampling point, history of COVID-19 episode, COVID-19-like illness, SARS-CoV-2 PCR testing, and close contact with COVID-19 patients after the preceding sampling point was enquired.

### Measurement of antibody titres

Antibody titres to SARS-CoV-2 spike protein (S-IgG) and neutralising antibody titres of all collected serum samples were measured using three commercially available chemiluminescence enzyme immunoassay (CLEIA) methodology-based reagents. First, IgG antibody titres against the SARS-CoV-2 spike protein S1 subunit receptor-binding domain (RBD) were measured using SARS-CoV-2 IgG II Quant reagents(Abbott Laboratories, Abbott Park, IL, USA) and an Alinity Analyzer (Abbott Laboratories, Abbott Park, IL, USA) according to the manufacturer’s instructions (Alinity RBD-IgG). Second, the IgG antibody titres against the anti-SARS-CoV-2 spike protein were measured using other chemiluminescence enzyme immunoassay (CLEIA) methodology-based systems and reagents: HISCL Analyzer (Sysmex Corporation, Kobe, Japan) and HISCL SARS-CoV-2 S-IgG reagents (Sysmex Corporation, Kobe, Japan) according to the manufacturer’s instructions (HISCL S-IgG). Finally, the neutralising ability of SARS-CoV-2 was assessed using the STACIA Analyzer (LSI Medience Corporation, Tokyo, Japan) and STACIA SARS-CoV-2 Neutralization Antibody Test (MBL Corporation, Nagoya, Japan), which assess neutralising ability by measuring the inhibitory activity of ACE 2 enzyme and RBD coated beads (STACIA Neut-Ab). Titres under the lower limit of detection (0.1 U/mL) were treated as 0.1 U/mL.

### Measurement of cellular immunity

T-cell immunity against SARS-CoV-2 was assessed using an interferon gamma releasing assay (IGRA). Based on the limited reagents supply, only the first 600 participants were selected at the time of the first sample collection, and whole blood samples were collected in lithium heparin tubes at the first, third, and final sampling points. Samples were transferred to four QuantiFERON SARS-CoV-2 tubes (Qiagen, Hilden, Germany): coated with antigen 1, antigen 2, phytohemagglutinin (positive control), and no peptide (negative control). Antigen 1 is an epitope of CD4+ T cells derived from the S1 subunit, and antigen 2 is an epitope of CD4+ and CD8+ T cells derived from the S1 and S2 subunits. The tubes were incubated at 37°C for 16–24 hours and enzyme-linked immunosorbent assays were performed using the QuantiFERON SARS-CoV-2 ELISA kit (Qiagen, Hilden, Germany) according to the package insert using an AP-96 auto microplate enzyme immunoassay reader (Kyowa Medex, Tokyo, Japan), which were quality controlled daily. The dynamics of interferon gamma levels for antigen 1 (IFN for Ag1) and antigen 2 (IFN for Ag2) after correction for negative control were investigated.

### Detection of suspected breakthrough infections among participants

In order to detect all the breakthrough infections including asymptomatic or undiagnosed cases during the study period, we measured the IgG antibody titre against the nucleocapsid protein of SARS-CoV-2, which is not affected by the vaccine which codes for the spike protein regions of SARS-CoV-2. For measuring the immunoglobulin G (IgG) antibody titre against the nucleocapsid protein (N-IgG), we used the HISCL Analyzer (Sysmex Corporation, Kobe, Japan) and HISCL SARS-CoV-2 N-IgG reagents (Sysmex Corporation, Kobe, Japan) according to the manufacturer’s instructions. We defined suspected breakthrough infections as follows: N-IgG test was negative before and after vaccination and turned positive during the 6 months after vaccination. N-IgG test positivity was determined according to the manufacturer’s cut-off (10 SU/mL). Clinical backgrounds and S-IgG, neutralising antibody titres, and QuantiFERON measurement of sampling points before breakthrough infection were compared between participants with or without suspected breakthrough infection.

### Statistical analysis

Participants who did not receive two doses of vaccination and those who did not who did not complete 5 points of sample collection were excluded from the statistical analysis set.

Summary statistics of the participants were constructed using frequencies and proportions for categorical data and mean and standard deviation (SD) for continuous variables. We compared antibody titres and interferon gamma levels of each sample collection point using paired t-test. A comparison between participants with or without suspected breakthrough infection was performed using Fisher exact tests categorical values, and Mann-Whitney U tests for continuous values according to the small number of suspected breakthrough infection cases. All statistical analyses were performed using JMP, version 15 and SAS software, version 9.4 (SAS Institute, Cary, NC, USA). Statistical significance was set at p < 0.05.

## Results

Out of the 673 healthcare workers and university staff members who were included in the present study, 3 participants who did not receive two doses of vaccination and 62 participants who did not complete 5 points of sample collection were eliminated. A total of 608 participants were included in the analysis (Table 1).

**Table 1.**
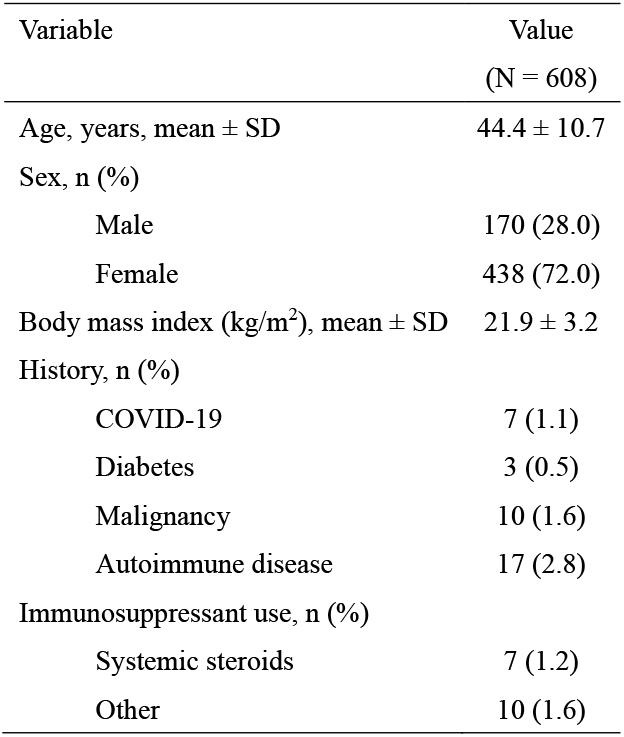
Demographics of the study participants

The antibody titres before vaccination were 9.8 ± 109.1 AU/mL in Alinity RBD-IgG, 0.2 ± 3.6 SU/mL in HISCL S-IgG and 1.0 ± 0.6 U/mL in STACIA Neut-Ab. After 3 weeks of vaccination the antibody titres increased to 15,443.5 ± 9,655.2 AU/mL in Alinity RBD-IgG, 406.0 ± 242.7 SU/mL in HISCL S-IgG, and 23.6 ± 14.1 U/mL in STACIA Neut-Ab. Antibody titres waned with time after vaccination and that after 6 months from vaccination were 1,576.8 ± 5080.2 AU/mL in Alinity RBD-IgG, 63.9 ± 195.9 SU/mL in HISCL S-IgG, and 3.3 ± 4.9 U/mL in STACIA Neut-Ab (Fig 1). Correlation analysis among the Alinity RBD-IgG, HISCL S-IgG, and STACIA Neut-Ab test results of all samples collected in the study demonstrated sufficiently high correlations (Supplement Fig S1).

**Fig. 1.**
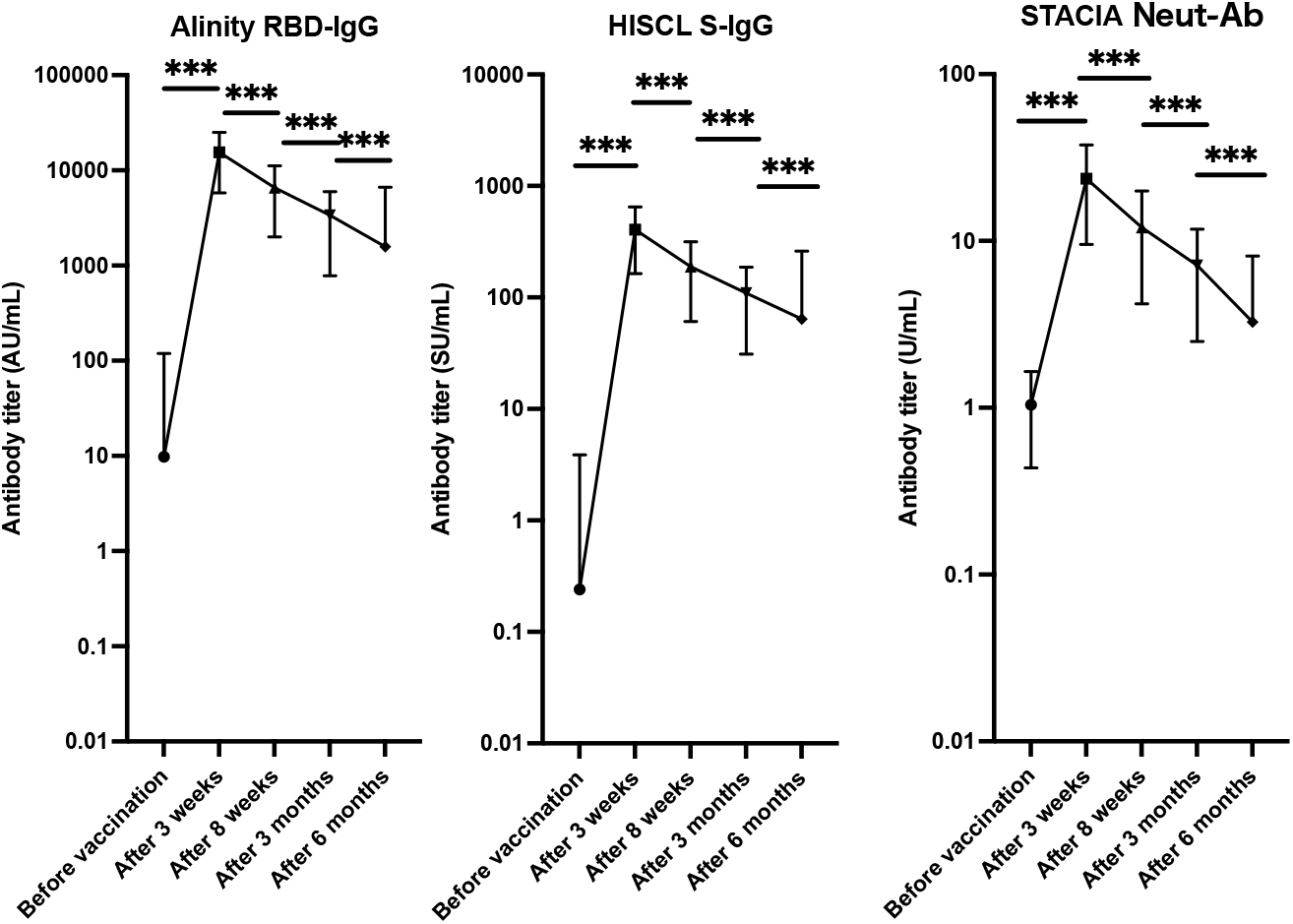
Dynamics of the anti-SARS-CoV-2 antibody response after vaccination with two doses of the BNT162b2 vaccine The plot shows the dynamics of the mean antibody titre at the five sample-collection time-points. The error bars indicate the standard deviation. Alinity RBD-IgG; antibody titre for the receptor-binding domain of SARS-CoV-2 was measured using SARS-CoV-2 IgG II Quant reagents and Alinity System (Abbott Laboratories, Illinois, USA), HISCL S-IgG: antibody titre for anti-SARS-CoV-2 spike protein measured using HISCL SARS-CoV-2 S-IgG reagents and an HISCL Analyzer (Sysmex Corporation, Kobe, Japan); STACIA Neut-Ab: neutralising ability for SARS-CoV-2 using STACIA SARS-CoV-2 Neutralization Antibody Test reagents (MBL Corporation, Nagoya, Japan), and a STACIA Analyzer (LSI Medience Corporation, Tokyo, Japan). Paired t-tests were used to calculate the p values (*** p<0.001). The Y-axes are on a logarithmic scale.

The QuantiFERON SARS-CoV-2 assay results demonstrated similar dynamics. Before vaccination, the assay was performed for 536 participants and IFN for Ag1 was 0.00 ± 0.08 IU/mL and IFN for Ag2 was 0.01 ± 0.07 IU/mL. After 8 weeks, IFN for Ag1 and IFN for Ag2 increased to 0.66 ± 0.87 IU/mL and 1.06 ± 1.25 IU/mL, respectively. However, IFN for Ag1 and IFN for Ag2 decreased to 0.37 ± 0.63 IU/mL and 0.62 ± 0.99 IU/mL, respectively, within 6 months after vaccination (Fig 2). The mean N-IgG titre was 0.6 ± 9.5 SU/mL before vaccination. The titre was not affected by the vaccination: 0.7 ± 8.4 SU/mL, 0.4 ± 4.7 SU/mL, and 0.3 ± 3.5 SU/mL after 3 weeks, 8 weeks, and 3 months of vaccination, respectively (Supplement Fig S2). After 6 months of vaccination, mean titre was 0.9 ± 8.9 SU/mL; however, six participants turned positive serologically and had suspected breakthrough infection.

**Fig. 2.**
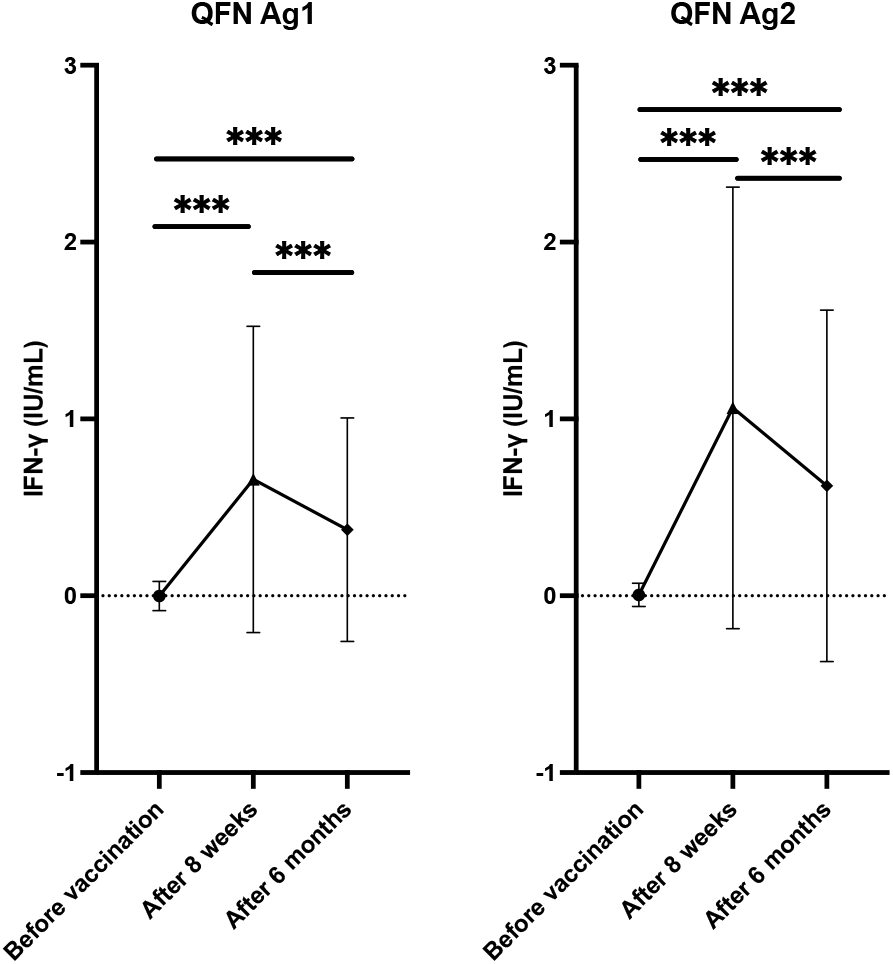
Dynamics of changes in cellular immunity after vaccination with two doses of the BNT162b2 vaccine The plot shows the dynamics of the mean interferon gamma levels in the blood stimulated by antigen 1 and 2 (IFN for Ag1 and IFN for Ag2) of QuantiFERON SARS-CoV-2 tubes (Qiagen, Hilden, Germany). The error bars indicate the standard deviation. Paired t-tests were used to calculate the p values (*** p<0.001).

All six participants with suspected breakthrough infection were female, without pre-existing illnesses or immunosuppressant use. One participant was diagnosed with COVID-19 according to the PCR test results performed 4 months after vaccination. One participant had close contact with COVID-19 patients after 5 months from vaccination; however, the PCR test result was negative. The other four participants were asymptomatic and had no history of close contact with patients with COVID-19 (Table 2). Compared with participants without suspected breakthrough, antibody titres before the breakthrough (after 3 month of vaccination) were equivalent, but titres after the breakthrough (after 6 month of vaccination) were significantly higher. However, the QuantiFERON results were not affected even after breakthrough (Table 3).

**Table 2.** Antibody titres in participants with suspected breakthrough SARS-CoV-2 infection

| Age<br>(years) | Sex | Antibody titre 3 months<br>after vaccination |  | Antibody titre 6 months<br>after vaccination |  | History of possible SARS-CoV-2 infection 3 to 6 months after<br>vaccination<sup>†</sup> |  |  |  |
| --- | --- | --- | --- | --- | --- | --- | --- | --- | --- |
|  |  | STACIA |  | STACIA |  | COVID-19 | COVID-19-like illness | SARS-CoV-2<br>PCR testing | Close contact<br>with COVID-19<br>patients |
|  |  | Neut-Ab,<br>U/mL | N-IgG,<br>SU/mL | Neut-Ab,<br>U/mL | N-IgG,<br>SU/mL |  |  |  |  |
| 40's | F | 6.45 | 0.5 | 51.38 | 19.8 | — | — | — | — |
| 20's | F | 9.86 | 0.1 | 52.92 | 22.3 | — | — | + | — |
| 40's | F | 10.73 | 0 | 42.21 | 79.9 | — | — | — | — |
| 30's | F | 9.92 | 5.4 | 20.55 | 173.3 | — | — | — | — |
| 50's | F | 7.68 | 0.1 | 53.73 | 55.3 | + | + | + | + |
| 40's | F | 5.17 | 0.1 | 53.24 | 83.0 | — | — | + | + |

<sup>†</sup> Based on answers provided in the questionnaire.
COVID-19, coronavirus disease; N-IgG, immunoglobulin G antibody against the SARS-CoV-2 nucleocapsid antigen; Neut-Ab, neutralisation antibody test; PCR, polymerase chain reaction; SARS-CoV-2, severe acute respiratory syndrome coronavirus 2

**Table 3.**
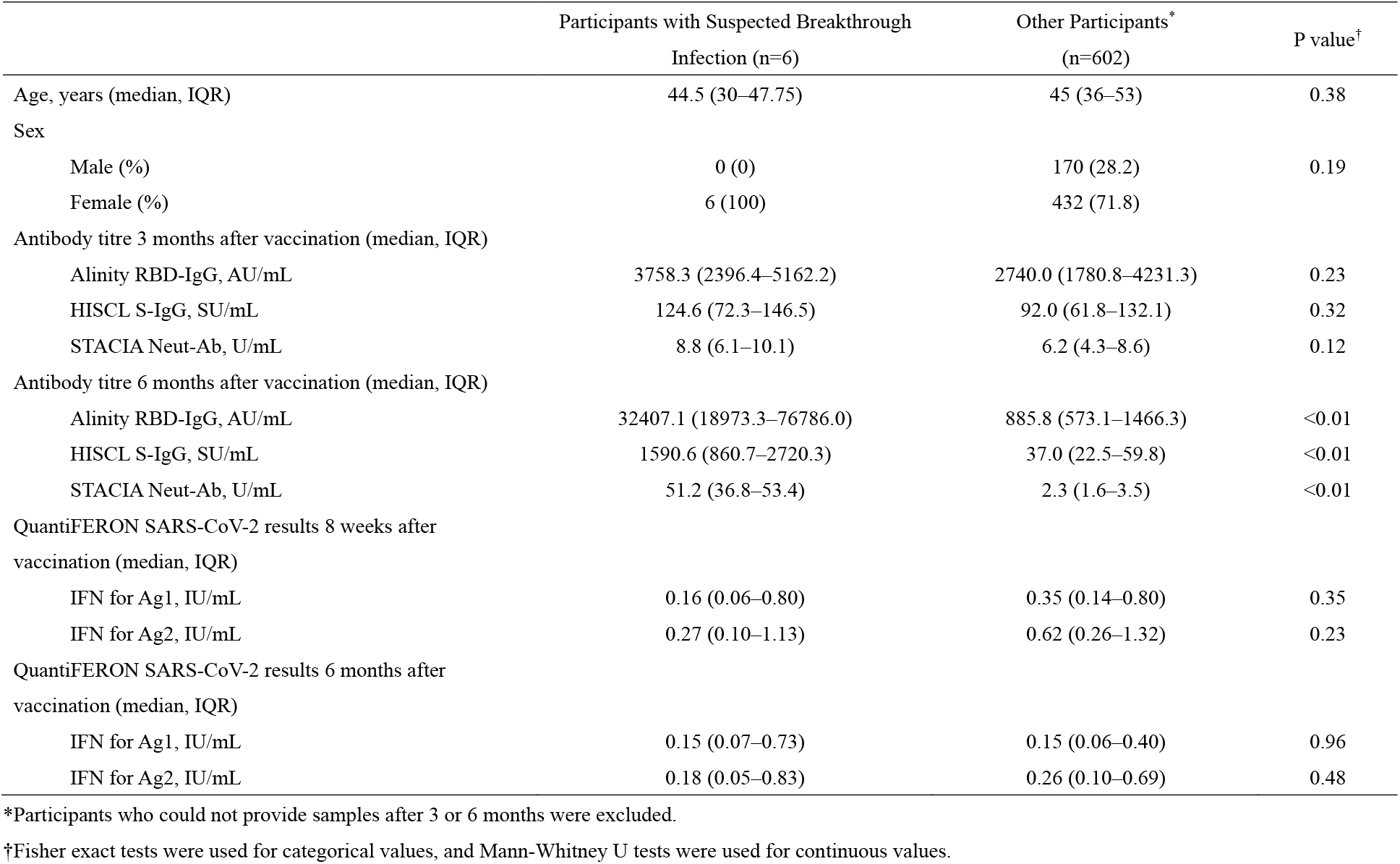
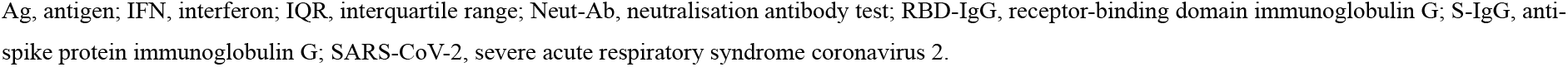
Difference in antibody titres and QuantiFERON results in participants with suspected breakthrough infections compared with the other participants

|  | Participants with Suspected Breakthrough<br>Infection (n=6) | Other Participants*<br>(n=602) | P value† |
| --- | --- | --- | --- |
| Age, years (median, IQR) | 44.5 (30–47.75) | 45 (36–53) | 0.38 |
| Sex |  |  |  |
| Male (%) | 0 (0) | 170 (28.2) | 0.19 |
| Female (%) | 6 (100) | 432 (71.8) |  |
| Antibody titre 3 months after vaccination (median, IQR) |  |  |  |
| Alinity RBD-IgG, AU/mL | 3758.3 (2396.4–5162.2) | 2740.0 (1780.8–4231.3) | 0.23 |
| HISCL S-IgG, SU/mL | 124.6 (72.3–146.5) | 92.0 (61.8–132.1) | 0.32 |
| STACIA Neut-Ab, U/mL | 8.8 (6.1–10.1) | 6.2 (4.3–8.6) | 0.12 |
| Antibody titre 6 months after vaccination (median, IQR) |  |  |  |
| Alinity RBD-IgG, AU/mL | 32407.1 (18973.3–76786.0) | 885.8 (573.1–1466.3) | <0.01 |
| HISCL S-IgG, SU/mL | 1590.6 (860.7–2720.3) | 37.0 (22.5–59.8) | <0.01 |
| STACIA Neut-Ab, U/mL | 51.2 (36.8–53.4) | 2.3 (1.6–3.5) | <0.01 |
| QuantiFERON SARS-CoV-2 results 8 weeks after<br>vaccination (median, IQR) |  |  |  |
| IFN for Ag1, IU/mL | 0.16 (0.06–0.80) | 0.35 (0.14–0.80) | 0.35 |
| IFN for Ag2, IU/mL | 0.27 (0.10–1.13) | 0.62 (0.26–1.32) | 0.23 |
| QuantiFERON SARS-CoV-2 results 6 months after<br>vaccination (median, IQR) |  |  |  |
| IFN for Ag1, IU/mL | 0.15 (0.07–0.73) | 0.15 (0.06–0.40) | 0.96 |
| IFN for Ag2, IU/mL | 0.18 (0.05–0.83) | 0.26 (0.10–0.69) | 0.48 |

\*Participants who could not provide samples after 3 or 6 months were excluded.
†Fisher exact tests were used for categorical values, and Mann-Whitney U tests were used for continuous values.
Ag, antigen; IFN, interferon; IQR, interquartile range; Neut-Ab, neutralisation antibody test; RBD-IgG, receptor-binding domain immunoglobulin G; S-IgG, anti-spike protein immunoglobulin G; SARS-CoV-2, severe acute respiratory syndrome coronavirus 2.

## Discussion

Our prospective cohort study including approximately 600 vaccinees revealed a decline in humoral and cellular immunity within 6 months after two doses of BNT162b2 vaccination by evaluating antibody titres and antigen-specific IFN-induction. This supports reports on waning effectiveness after the first two vaccine doses, suggesting the importance of the urgent booster dose to induce stronger immunity against SARS-CoV-2 [8–10].

Three commercially available CLEIA-based SARS-CoV-2 antibody measurement kits were used. The IgG antibody titres against RBD or S1 peptides were well correlated with neutralising antibody levels based on their ability to inhibit the formation of the complex of RBD and ACE2. Thus, measurement of anti-RBD or anti-S1 IgG titres might be substituted for the measurement of antibody activity to neutralize SARS-CoV-2 infection.

As most breakthrough infections are reported to be mild or asymptomatic, opportunities to improve diagnostic performance are limited [11]. The measurement of N-IgG made it possible to detect five asymptomatic breakthrough infections in the present study. One of them was subjected to PCR testing, but the result was negative. Although the negativity may be attributed to the sensitivity of PCR tests, it is plausible that a faster mean rate of viral load decline in fully vaccinated individuals than in unvaccinated individuals might give rise to a negative result [12].

The median antibody titre of participants with suspected breakthrough 3 months after vaccination was not lower than that of those without breakthrough infections. This suggests that breakthrough infections may occur not only in poor responders but also in good responders several months after BNT162b2 vaccination. Otherwise, the median antibody titre using STACIA Neut-Ab after 6 months of vaccination (51.2 U/mL) being higher than that after 3 weeks of vaccination (31.9 U/mL) seems to provide evidence that asymptomatic breakthrough infections exert a sufficient booster effect, consistent with observations previously obtained from breakthrough infections confirmed by PCR tests [13].

Notably, the IGRA results were not affected by breakthrough infections in the present study. This is similar to a report by Kato et al. [4] on discrepant antibody titres and IGRA results due to breakthrough infection. As the reason for the discrepancy between the antibody titres and the IGRA results has not been determined, further studies are warranted.

This study had several limitations. First, STACIA Neut-Ab, which was used as the neutralisation assay, could only detect the antibody activity to inhibit RBD binding to human ACE 2. Although the STACIA Neut-Ab results for monoclonal antibodies correlated well with the neutralisation assay results using in vitro SARS-CoV-2 infection in cells, neutralisation is thought to incompletely represent in vivo humoral immunity against SARS-CoV-2 with polyclonality [14]. Therefore, the verification of neutralising antibody using SARS-CoV-2 infected cells or animals is ideal. Second, cross-reactivity of the N-IgG antibody test has been reported. False positive results might be obtained due to cross-reactivity to coronavirus NL63 and 229E infections; therefore, N-IgG-positive conversion do not always signify breakthrough SARS-CoV-2 infection. However, the occurrence of NL63 and 229E infections cannot explain the elevation of HISCL S-IgG titres after 6 months of vaccination because of the specificity of the measured antibody to SARS-CoV-2 infections [15].

Third, the incidence of breakthrough infections could not be explained simply by waning humoral and cellular immunity. Occupational contact with COVID-19 patients, social activity, and prevalence of SARS-CoV-2 infection may have affected the results. Especially, between the fourth to fifth sample collection time-points, there was a high incidence of SARS-CoV-2 recorded in Japan during the Olympic and Paralympic games in Tokyo. Therefore, further investigation including more number and variety of participants is required to make firm conclusions about the relationship between immunity acquired by vaccination and breakthrough infections. Finally, in Japan, alpha variant infections were dominant during the third (winter, 2021) and fourth (spring, 2021) waves of the SARS-CoV-2 epidemic, and infections due to the delta variant were dominant during the fifth (summer, 2021) wave of the SARS-CoV-2 epidemic. Therefore, the rate of suspected breakthrough infections in our study was under the circumstance of an epidemic of delta variants. Currently, the omicron variant, which can escape the immunity of BNT162b2 vaccination [16], is dominant worldwide, including in Japan. Thus, the incidence of breakthrough infections may have increased. In conclusion, waning of humoral and cellular immunity within 6 months after two doses of BNT162b2 vaccine among Japanese healthcare workers and the occurrence of asymptomatic breakthrough infection was suspected in around 1 of 100 vaccinees. This information may aid in optimising vaccination to overcome the COVID-19 pandemic.

## Data Availability

All data produced in the present study are available upon reasonable request to the authors

