## Supplement Fig S1, Supplement Fig S2 for "Dynamics of antibody responses, cellular immunity, and breakthrough infections among Japanese healthcare workers during the 6 months after receiving two doses of BNT162b2 mRNA vaccine"

Supplement files

Fig S1. Correlations between 3 types of antibody test reagents

1)STACIA Neut-Ab vs Alinity RBD-IgG

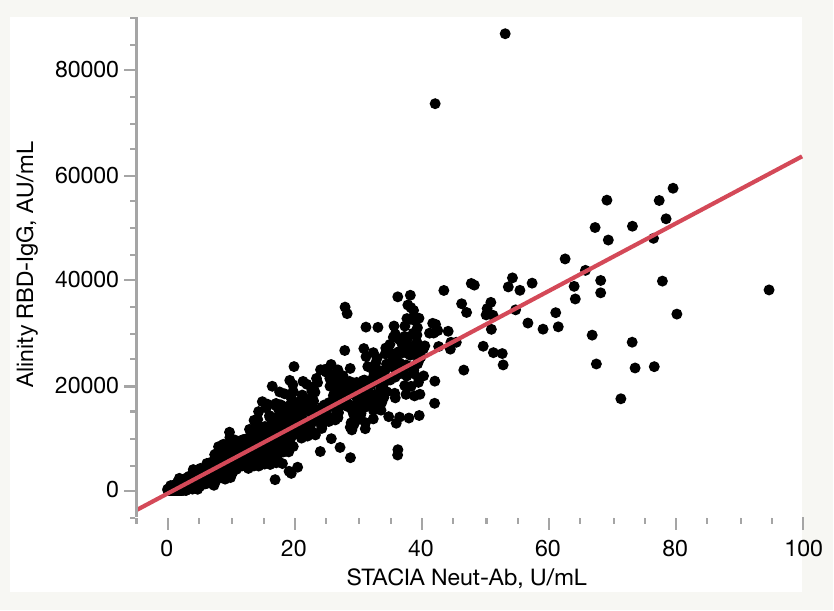

R^2^ = 0.89 (p < 0.01)

2)STACIA Neut-Ab vs HISCL S-IgG

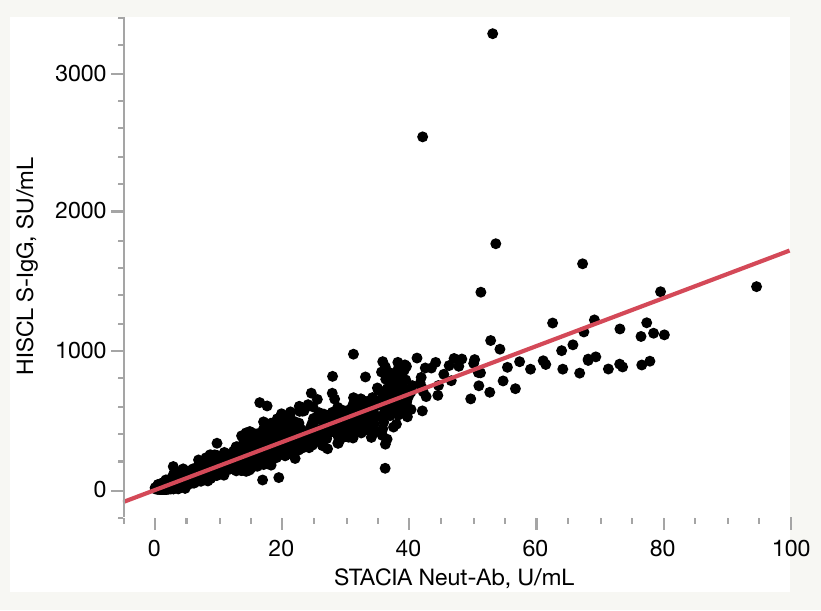

R^2^ = 0.87 (p < 0.01)

Fig S2. Dynamics of N-IgG titers.

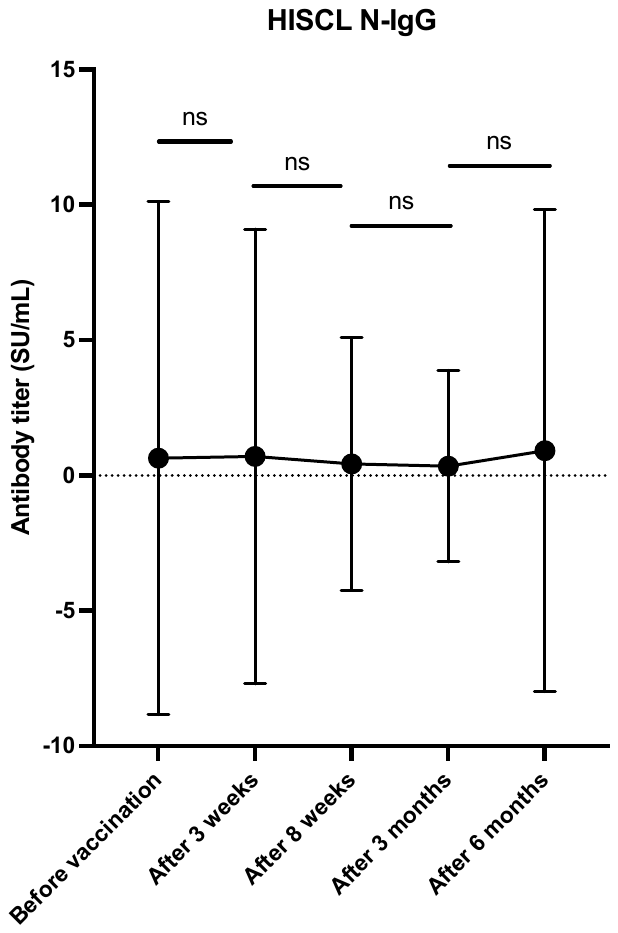

*Mean and standard deviations were demonstrated. Paired t-test were performed for statistical analysis.
